# Web-based education on Metabolism and Obesity is associated with improved lifestyle and health behaviours among Brazilian school teachers

**DOI:** 10.64898/2026.06.09.26355322

**Authors:** Paulo Henrique Evangelista-Silva, Camille Perella Coutinho, Camila Caldas Martins Correia, Talles Moraes, Ana Flávia Fernandes Ferreira, Ayumi Cristina Medeiros Komino, João Kleber Neves Ramos, Leila Affini, Renata Ligia de Araujo Furlan, Carla Roberta de Oliveira Carvalho

**Author notes:** Corresponding authors: /. Contributed equally (Paulo Henrique Evangelista-Silva, Camille Perella Coutinho, Camila Caldas Martins Correia).

## Abstract

**Background:** Obesity is a major global public health challenge, and teachers play a critical role in school-based health promotion. This study examined the perceived impact of a web-based educational program on metabolism and obesity delivered to Brazilian school teachers.

**Methods:** This analytical cross-sectional study included 217 teachers who responded to the evaluation questionnaire after attending the course between 2017 and 2022. Statistical analyses included logistic regression and chi-square tests.

**Findings:** Course completion rate was 81.98%, substantially exceeding the 5-15% typical of global MOOCs. However, ethnic disparities were observed: White respondents were 4.95 times more likely to complete the course than Black respondents (*p*=0.00097) and Brown respondents were 3.05 times more likely (*p*=0.0268) than Black respondents. Among non-completers, lack of time (64.7%) was the primary barrier. Participation was concentrated in São Paulo (77%), with no respondents from three northern states. Perceived difficulty showed a non-significant trend (*p*=0.0893) whereby Black respondents had the lowest predicted difficulty; the most challenging course material was Scientific Content/Reading papers (50%). Completion was strongly associated with applying learned activities in teaching (*p*<2.2×10⁻¹⁶); 57.1% of completers implemented health-promoting activities, most commonly games, healthy eating, and combined diet-physical activity habits. Completers also reported significant improvements in lifestyle decisions (*p*=1.76×10⁻²¹) and healthy habits (*p*=9.35×10⁻⁷⁷), including better diet and increased physical activity.

**Conclusion and interpretation:** Course completion was associated with reported improvements in diet, physical activity, nutrition knowledge, and teaching practices among Brazilian teachers. However, marked ethnic and regional disparities were observed. White teachers were nearly five times more likely to complete the course than Black teachers. The absence of respondents from northern states reveals that scalability without equity widens existing gaps. Online access to free courses alone does not democratize education; mitigating ethnic and regional disparities must be a priority for digital health interventions.

## 1 INTRODUCTION

Obesity represents one of the most critical and complex global public health challenges of the 21st century, contributing substantially to the increasing burden of non-communicable diseases (NCDs), including type 2 diabetes, cardiovascular diseases, and cancer^1^. Increasingly, obesity has been conceptualised as part of a broader global syndemic, interacting with undernutrition and climate change to exacerbate both human and planetary health risks^2^. According to the World Health Organization (WHO), obesity is defined as abnormal or excessive fat accumulation that impairs health and arises from a complex interplay of genetic, epigenetic, environmental, and behavioural factors ^3,4^.

The prevalence of obesity has increased dramatically worldwide, affecting all age groups, with particularly concerning trends among children and adolescents^5^. In Brazil, approximately 13% of children aged 5–9 years are obese, reflecting early exposure to obesogenic environments^6,7^. Early-life determinants including maternal nutritional status, infant feeding practices, and intergenerational influences play a critical role in shaping long-term metabolic outcomes^8,9^. Childhood obesity is also associated with immediate physiological consequences, including altered growth patterns, accelerated bone maturation, early pubertal onset, and endocrine disturbances^10–12^.

Given the multifactorial nature of obesity, effective prevention strategies must extend beyond clinical approaches and incorporate behavioural and educational interventions. In this context, schools are widely recognised as strategic environments for health promotion, with the potential to influence behaviours during critical stages of development. Teachers play a central role in this process, acting as mediators of knowledge, role models, and facilitators of behavioural change^13,14^. Evidence shows that school-based interventions delivered by trained teachers can reduce body mass index trajectories and the incidence of overweight in children ^15^, as well as improve students’ nutritional knowledge and health-related behaviours ^16,17^.

To address these challenges and gaps, the extension course: *“Metabolism and Obesity: a physiological approach”* was developed as a web-based educational intervention aimed at training Brazilian school teachers in key concepts related to metabolism, obesity, and healthy lifestyle promotion. Since its implementation in 2017, the course has reached educators across multiple regions of Brazil; however, its impact on lifestyle behaviours, teaching practices, and dissemination of knowledge within school settings has not been systematically evaluated.

Therefore, the present study aimed to evaluate the impact of the web-based course “*Metabolism and Obesity: a physiological approach*” on Brazilian school teachers. Specifically, the study sought to: (i) assess changes in lifestyle and health behaviours following course participation; (ii) examine the extent to which acquired knowledge was translated into pedagogical practices and school-based activities; and (iii) evaluate the effectiveness of the intervention as a scalable strategy for health promotion within the educational system. The findings indicate that course completion led to improvements in both teachers’ personal health behaviours and classroom practices, while also revealing significant ethnic and regional inequities.

## 2 METHODS

### 2.1 Study design and setting

This cross-sectional analytical study evaluated the impact of the online educational course “*Metabolism and Obesity: a physiological approach*” on teachers in the Brazilian public school system. The study consisted of two main components: (i) the development and implementation of the online course and (ii) a post-intervention evaluation using a structured questionnaire. The course was delivered nationwide between 2017 and 2022 and made available through the University of São Paulo’s extension platform (Moodle: https://cursosextensao.usp.br/course).

### 2.2 Intervention: course development and structure

The continuing education program “*Metabolism and Obesity: a physiological approach*” was established in 2017 by researchers from the Human Physiology Graduate Program at the Institute of Biomedical Sciences, University of São Paulo (ICB-USP). The course was conceived as a short-duration, distance learning (EAD) university extension initiative, designed to disseminate scientific knowledge to the external community, specifically primary and secondary school teachers from public and private schools across Brazil. Beyond updating teachers on metabolism and obesity, the course also contributes to the technical and humanistic training of graduate and undergraduate students from the Department of Physiology and Biophysics, which hosts the program.

The Department of Physiology and Biophysics at ICB-USP has a recognized tradition in continuing education for schoolteachers. Between 2005 and 2013, the department offered the “Winter Course in Physiology,” an intensive 40-hour, week-long in-person program held during school recess. Café-Mendes et al., (2016)^38^ thoroughly described this initiative, demonstrating that it successfully updated secondary school teachers in various physiology topics while providing graduate students with valuable teaching experience. That course was highly regarded by participants and served as an important precursor for subsequent educational outreach initiatives ^38^. Building directly on this foundation, the *Metabolism and Obesity* course, also known informally as the “Winter Course” in its online format, was launched in 2017 as a fully remote evolution of the original proposal. Unlike its in-person predecessor, which required teachers to travel to São Paulo for one week, the new format adopted a self-paced, distance learning model to overcome geographical and time constraints, enabling nationwide reach and flexible participation.

The initial motivation for creating a course with this specific theme stemmed from the preclinical research experience of the department’s faculty and aligned with the World Health Organization’s “Beat Diabetes” campaign launched in 2016. The course curriculum consists of six video lecture modules, recorded primarily by graduate students from the Human Physiology Graduate Program (ICB). These modules are supported by supplementary texts and self-assessment questions, and all materials were developed under the supervision of at least two faculty members from the Department of Physiology and Biophysics. Designed to bridge the gap between bench science and school-based health education, the curriculum covers the physiological continuum from nutrient absorption to the epigenetic determinants of metabolic syndrome (**Figure 1**).

**Figure 1:**
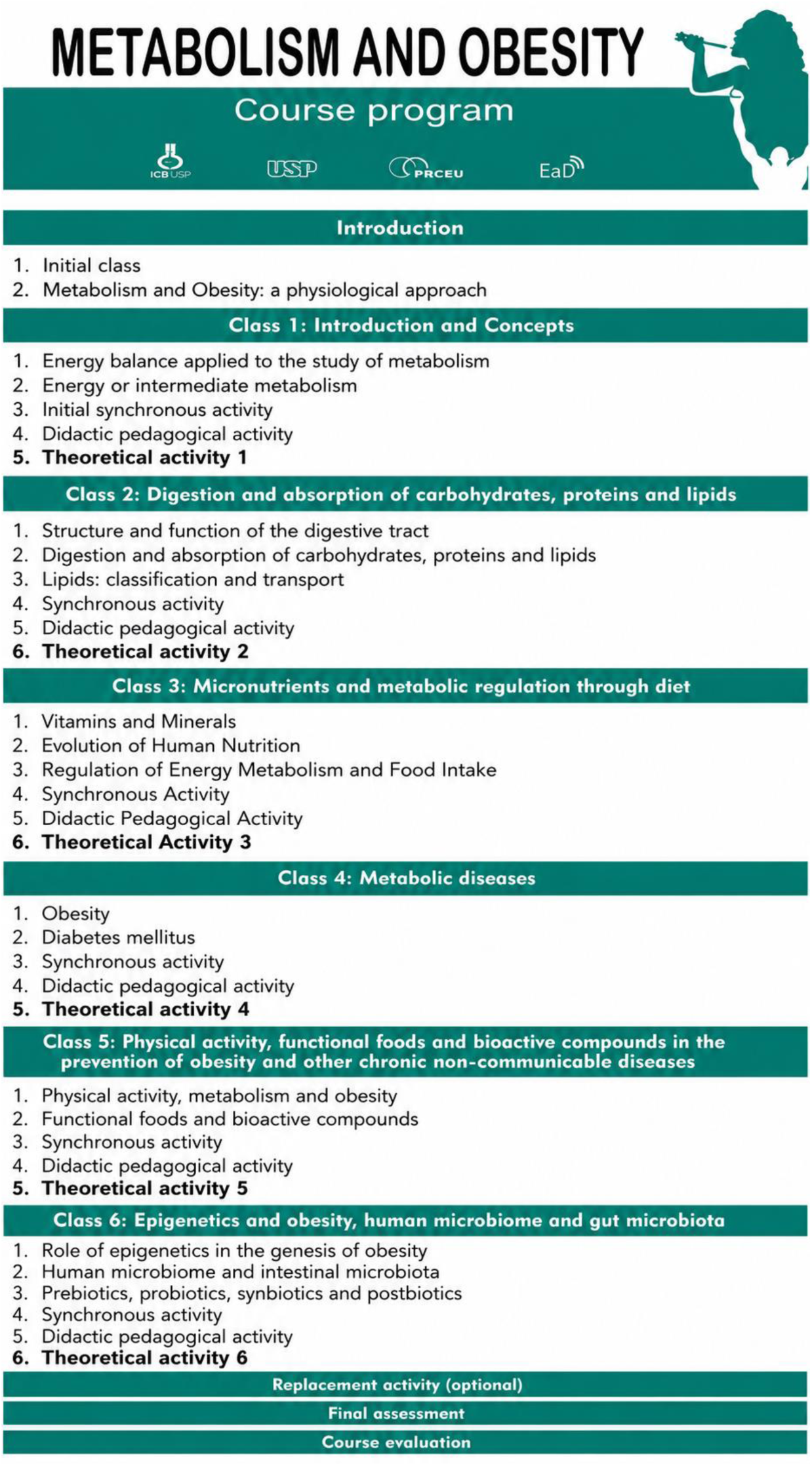
Curricular framework of the continuing education program on Metabolism and Obesity, detailing thematic modules and their integration with translational pedagogical activities.

In 2019, the “*Bio Cientista Mirim*” (Little Scientist Biologist) University Extension Project team joined the course, developing exclusive paradidactic materials aligned with the Brazilian National Common Curricular Base (BNCC) for enrolled teachers. To ensure effective translation of scientific evidence into pedagogical practice, each module is structured around three core pillars: (i) Evidence-Based Theory, featuring high-level lectures on the pathophysiology of chronic non-communicable diseases; (ii) Knowledge Consolidation, using theoretical activities as self-assessment milestones; and (iii) Translational Pedagogy, implemented through the *Bio Cientista Mirim* initiative. This framework provides educators with a streamlined suite of validated tools for classroom implementation, integrating simplified laboratory protocols to demonstrate metabolic processes with inquiry-based learning strategies and educational games designed to enhance health literacy among adolescents. The initiative also includes structured scripts to facilitate critical discussions on lifestyle interventions and the prevention of obesity-related comorbidities.

Since 2020, the course has been updated annually. During the COVID-19 pandemic, the model was refined to include synchronous biweekly remote meetings with participants, an important forum for presenting dynamics and activities promoted by *Bio Cientista Mirim*, as well as for discussions and activity feedback throughout each module. In addition, computer-based learning games have been developed to support content retention and differentiated assessment. Participants are evaluated each module through questionnaires and forums, where doubts and discussions can be addressed throughout the course.

The course has been consistently praised by participants and has fulfilled its role of disseminating knowledge to the external community across Brazil, while strengthening student training within the university and providing access to the university extension pillar.

### 2.3 Participants and data collection

Eligible participants included teachers who attended the course between 2017 and 2022. A total of 357 former participants were invited via email to complete the evaluation questionnaire, of whom 217 responded and, thus, were included in the analysis. Participation was voluntary, anonymous, and without financial incentive. Informed consent was obtained from all participants prior to data collection. The study was approved by the Research Ethics Committee of the University of São Paulo (CAAE #59027922.9.0000.5467).

Data were collected using a structured online questionnaire administered via Google Forms (https://docs.google.com/forms/). The instrument was designed to capture multiple dimensions of the intervention, including sociodemographic characteristics, course experience, perceived difficulty, lifestyle-related changes, and the implementation of health-related activities in school and community settings.

### 2.4 Questionnaire

The questionnaire consisted of 24 items combining closed-ended and open-ended questions. Quantitative responses were primarily collected using Likert-type scales (1–5), allowing the assessment of perceived changes in lifestyle behaviours, decision-making, and pedagogical practices following course participation.

The instrument also included open-response fields to capture qualitative insights regarding participants’ experiences, application of course content, and suggestions for improvement. The full questionnaire structure is presented below (**Table 1**), detailing all variables collected and their respective response formats.

**Table 1:** Structure and content of the evaluation questionnaire administered to course participants.

|  |  |
| --- | --- |
| 1) What was your age group during the Course? | • Between 20 and 30 years<br>• Between 31 and 40 years<br>• Between 41 and 50 years<br>• Between 51 and 60 years<br>• Over 60 years |
| 2) Which ethnic group do you identify as? | • White<br>• Brown<br>• Black<br>• Indigenous<br>• Yellow<br>• White/Black<br>• Mixed<br>• None |
| 3) In which year were you a student of the Course? | 2017, 2018, 2019, 2020, 2021, 2022 |
| 4) What was your occupation when you took the Course? | (Open-ended response: Users type their answer) |
| 5) What type of education network did you work in when you took the course? | • Public Network<br>• Private Network<br>• Public and Private Network<br>• None |
| 6) In which State did you work when you took the Course? | • Alagoas (AL)<br>• Amapá (AP)<br>• Amazonas (AM)<br>• Bahia (BA)<br>• Ceará (CE)<br>• Distrito Federal (DF)<br>• Espírito Santo (ES)<br>• Goiás (GO) |
| 7) How did you hear about the Course? | • Friends, Facebook, Instagram, Website, Telegram, Television, Whatsapp, Email |
| 8) On a scale of 1 to 5, how difficult was it for you to enroll in the Course? | 1- Easy; 2- Moderate; 3- Challenging; 4- Difficult; 5- Very Difficult |
| 9) Would you like to leave us any suggestions to improve the experience of future users when it comes to registering for the Course? | (Open-ended response: Users type their answer) |
| 10) Have you completed the entire course? | • Yes or No |
| 10.1) Why didn't you complete the Course? | (Open-ended response: Users type their answer) |
| 10.2) Indicate the importance of the factors below in the decision to interrupt the Course? | • High<br>• Medium<br>• Low<br>• None |
| 10.3) Are you currently still working in the education sector? | • Yes, No or Never acted |
| 11) From 1 to 5, how compatible do you consider the relationship between the topics covered in the Course and current events? | (1- Incompatible; 2- Partially incompatible; 3- Neither incompatible nor compatible; 4- Partially compatible; 5- Compatible) |
| 12) Did you develop any activity that you learned during the Course with your students (students to whom you teach or taught classes)? | • Yes or No |
| 12.1) If your answer to the previous question was "Yes", then tell us about your experience. | (Open-ended response: Users type their answer) |
| 13) Did you develop any activities that you learned during the Course with your school community (other students, teachers, school staff and/or parents) and/or local community (for example, people in your neighbourhood)? | • Yes or No |
| 13.1) If your answer to the previous question was "Yes", then tell us about your experience. | (Open-ended response: Users type their answer) |
| 14) From 1 to 5, how much are topics about healthy lifestyle discussed in the books used in the classroom? | 1- Absolutely no mention of the subject; 2- Very slightly; 3- Neither much nor little; 4- There is considerable discussion (one paragraph/page); 5- Strongly discussed. There is an entire chapter or more devoted to the subject |
| 14.1) Still on the previous question: If you agreed that the topic of healthy lifestyle is discussed in the books, indicate here how they are addressed in the books (which topics, themes, etc.) | (Open-ended response: Users type their answer) |
| 15) From 1 to 5, please describe the impact of the Course on you considering: | (Open-ended response: Users type their answer) |
| A) Your decision-making regarding a healthier lifestyle after completing the Course. | 1- It got worse and worse; 2- It did not get better; 3- It did not get better, nor did it get worse; 4- It got a little better; 5- It got completely better |
| B) Changing your daily habits towards a healthier lifestyle after completing the Course. | 1- I did not even try; 2- I tried, but I could not change; 3- My habits were already good; 4- I tried and managed to change a little; 5- They changed completely for the better) |
| C) The change in your way of thinking about a healthy lifestyle, planning new activities and developing strategies at school/institution after completing the Course. | 1- I had never thought about the subject; 2- It did not change at all; 3- It changed partially; 4- It changed; 5- It changed completely, I developed critical thinking about the subject and I started planning new activities and strategies to apply at school |
| D) The change in your way of seeing the importance and need to work on topics about healthy eating and obesity with students in the schools or institutions where you work. | 1- I still thought the subject was unnecessary; 2- I made an effort and began to understand its importance; 3- I already knew its importance, but I didn't propose any activities for my students; 4- I started to treat the subject more seriously and plan activities for the students; 5- It changed completely; I started to realize the extent of my responsibility for the health of my students and, therefore, the need to work on these topics |
| E) The change in the way you approach content about healthy lifestyle in the classroom or environment/institution in which you work after completing the Course. | 1- I continued without covering any content; 2- I made an effort and started to apply the activities proposed in the book; 3- I continued with the same approach I had before the Course; 4- I added a little of what I learned in the Course, whether in the content of the classes or when planning new activities to be developed; 5- I completely innovated my approach when developing strategies on healthy lifestyle at school and/or in the community |
| 16) Indicate the healthy habits acquired after completing the Course: | (Open-ended response: Users type their answer) |
| 17) If you are currently teaching classes, do you work on any healthy lifestyle topics? | • Yes, No or I am not teaching classes currently |
| 18) Indicate the topic(s) about healthy lifestyle that you work on or have worked on in the classroom/institution where you work, with your students. If you do not work on it or have never worked on it, tell us why, or if you intend to, etc. | (Open-ended response: Users type their answer) |
| 19) Have you ever mentioned to the principal, teachers of other subjects, such as Physical Education, or the nutritionist or school lunch staff about having participated in this course? Were you able to promote any changes, such as offering healthier snacks to students, | (Open-ended response: Users type their answer) |
| increasing the frequency of physical activities, or even creating a vegetable garden at the school? |  |
| 20) From 1 to 5, to what extent did the synchronous activities (meetings with the distance learning course coordinators) contribute to your understanding of the content and/or encourage you to make healthier decisions? | 1- In fact, they were a hindrance and demotivating; 2- They can improve; 3- They were indifferent; 4- They added knowledge exchanges, they were motivating and just right; 5- I would like them to have more synchronous classes |
| 21) Would you have liked to have had synchronous activities with the Course coordinators, in addition to the recorded classes? | • Yes or No |
| 21.1) Comment on the reason for your choice | (Open-ended response: Users type their answer) |
| 22) Were you able to create any new activities for your students based on what you learned in the Course? | (Open-ended response: Users type their answer) |
| 22.1) If you answered "Yes" to the previous question, what activity was that? | (Open-ended response: Users type their answer) |
| 23) How difficult was it for you to learn the content taught in the Course? | 1- Easy; 2- Moderate; 3- Challenging; 4- Difficult; 5- Very Difficult |
| 23.1) If in the previous question you rated your level of difficulty as 4 or 5, comment on the content(s) you had the most difficulty learning. | (Open-ended response: Users type their answer) |
| 24) Are there any considerations regarding the Course or that you consider relevant that you would like to tell us about? Suggestions for improving the Course are also welcome. | (Open-ended response: Users type their answer) |

### 2.5 Outcomes

The primary outcomes were defined as self-reported changes in lifestyle and health behaviours following participation in the course, assessed using Likert-scale responses (1-5). These included changes in decision-making related to healthy living, adoption of healthier daily habits, and modifications in attitudes toward health promotion. A second primary outcome was the application of acquired knowledge in educational practice, measured by participants’ self-reported implementation of health-related activities in classroom and school settings (binary: yes/no).

Course completion (yes/no) was treated as both an outcome and an explanatory variable, given its central role in subsequent analyses examining its association with behavioural changes and pedagogical implementation. Perceived course difficulty was assessed as an additional outcome using ordinal categories (1-5 scale). Secondary outcomes included the type and frequency of health-related activities developed in schools, engagement with the broader school community, and qualitative reports describing the nature of implemented interventions. Sociodemographic variables (e.g., age and ethnicity) were considered explanatory variables in the analysis of course completion and perceived difficulty.

### 2.6 Statistical analysis

Data analysis was conducted to evaluate the relationship between demographic characteristics, course completion rates, and the application of activities learned during the course among participants. All analyses were performed using the R programming environment (version 4.3.3; https://www.R-project.org), with the RStudio interface. Descriptive statistics were used to summarize the demographic characteristics of the study population, with absolute and relative frequencies calculated for categorical variables such as ethnicity and course completion. The study population consisted of 217 respondents s from across Brazil, categorized as White (53.95%), Brown (31.63%), Black (12.56%), and Other (less than 1%), the latter including mixed-race and Asian individuals.

A logistic regression model was applied to examine the association between ethnicity and course completion, using the Black group as the reference category. Results were reported as odds ratios (OR) with 95% confidence intervals (CI) and corresponding p-values, with statistical significance defined as *p* < 0.05. To assess the association between course completion and the application of activities learned during the course, a chi-square test was performed using the chisq.test() function from the stats package in R. Additionally, the relationship between age, ethnicity, and perceived course difficulty was analyzed using logistic regression models, from which predicted probabilities of experiencing course difficulty were estimated and visualized across demographic groups. Data visualization was performed using the ggplot2 package in R.

## 3 RESULTS

### 3.1 Demographics and geographical distribution of the “*Metabolism and Obesity: a physiological approach*” course respondents

Between 2017 and 2022, 217 teachers enrolled in the Metabolism and Obesity course completed the evaluation questionnaire, constituting a nationwide sample from Brazil. However, participation was markedly uneven, with the state of São Paulo accounting for 77% of all respondents (**Figure 2A**). When analysed longitudinally, São Paulo was predominant from 2017 to 2021, consistently representing the majority of respondents each year (2017: 85%; 2018: 100.0%; 2019: 81%; 2020: 68%; 2021: 61%; 2022: 66%**) (Supplementary document 1)**. Year-stratified percentage calculations confirmed this pattern, whereas no registered respondents originated from the northern states of Acre, Roraima, or Amapá (**Figure 2A**). In the remaining years, respondents from other Brazilian states were also observed, including PR, ES, RJ, MG, AM, RS, MT, and others, indicating a gradual expansion of geographical reach over time.

**Figure 2:**
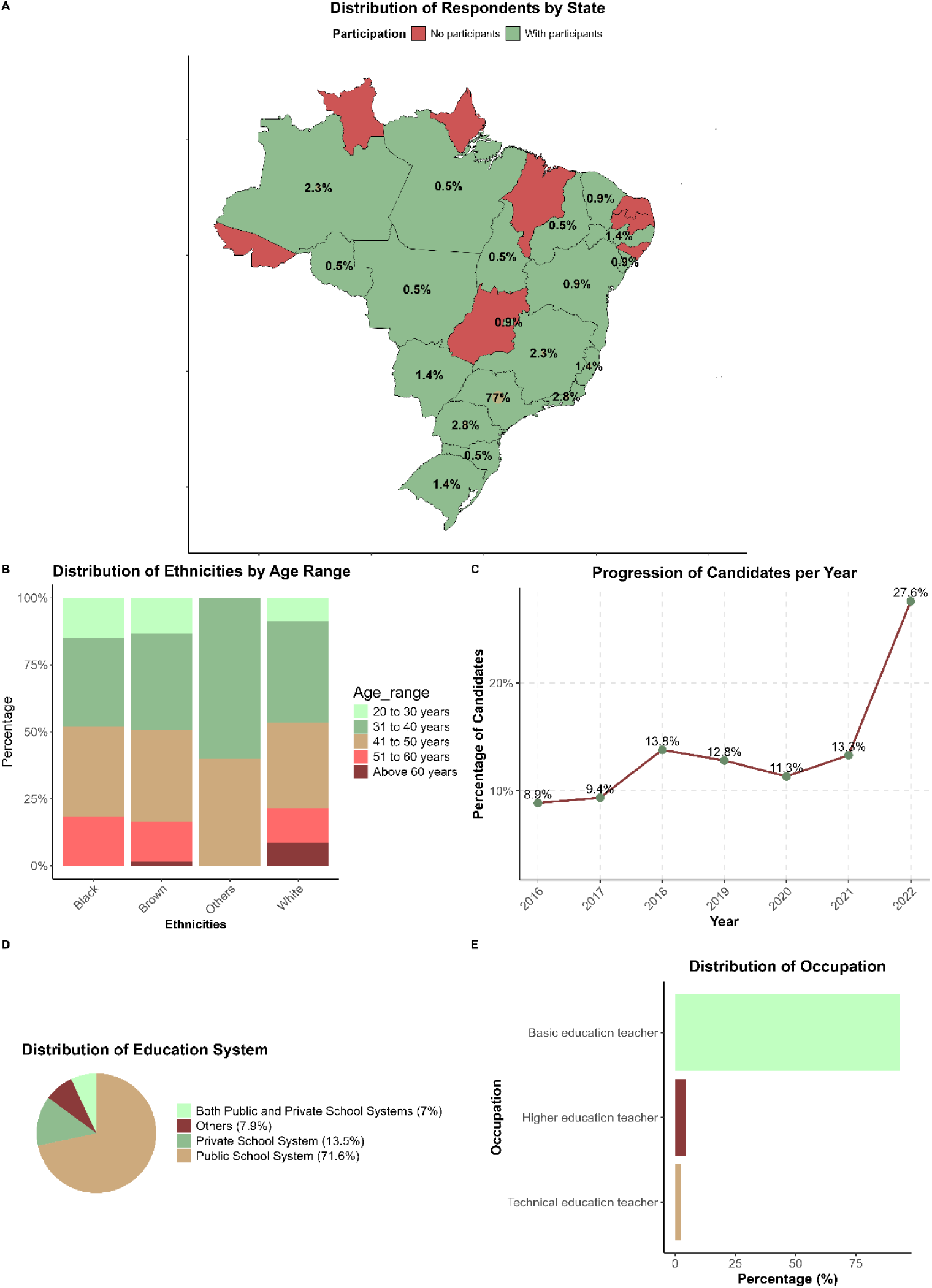
Demographic characteristics, geographical distribution, and professional background of respondents enrolled in the Metabolism and Obesity course between 2017 and 2022. **(A)** Geographical distribution of respondents across Brazilian states. **(B)** Distribution of respondents by age group and self-reported ethnicity. **(C)** Temporal evolution of course respondents from 2017 to 2022, with annual growth rates indicated. **(D**) Distribution of respondents by type of school. **(E)** Educational level in which respondents were engaged.

This distribution should be interpreted in light of the course dissemination strategy, as recruitment was primarily restricted to the state of São Paulo prior to the COVID-19 pandemic due to institutional collaboration and local professional incentives. Despite recruitment originally being planned only for the state of São Paulo, a few teachers from other states were included after directly contacting the organizing committee. Given this observed increase in interest, we decided to open the course nationwide. From 2020 onwards, the course was opened nationwide, resulting in increased participation from other regions. Therefore, the observed geographical disparity likely reflects both structural aspects of programme implementation and differences in regional access or engagement with online continuing education.

The demographic composition of the sample was predominantly White (53.95%), followed by Brown (31.63%) and Black (12.56%) responders. Individuals of Asian descent and other mixed-race backgrounds comprised less than 1% of the sample and were categorised as “Other” (**Figure 2B**). Age distribution was similar across ethnic groups, with the highest proportion of respondents in the 31-40-year age bracket, followed by those aged 20-30 years. Respondents older than 60 years were minimally represented, indicating that the course primarily reached early-career and mid-career educators rather than those approaching retirement (**Figure 2B**). Temporal analysis of enrolment data revealed a progressive increase in course participation over time, with a pronounced acceleration beginning in 2021 and a peak annual growth rate of 27.6% in 2022 (**Figure 2C**). This pattern probably reflects heightened visibility, accessibility, or demand for online educational initiatives during and after the COVID-19 pandemic.

Regarding professional characteristics, most respondents were employed in public schools, and the majority worked in basic education (**Figure 2D, 2E**). These findings underscore the course’s relevance to primary and secondary educational settings. Collectively, the results indicate that the intervention successfully engaged a population of active classroom teachers with direct influence on school-aged children and adolescents, albeit with notable regional and demographic imbalances that may limit generalisability to the entire Brazilian teaching workforce.

### 3.2 Ethnic disparities in course completion and perceived difficulty: who finishes and who struggles

Overall, 81.98% of respondents s successfully completed the Metabolism and Obesity course, whereas 18.02% did not finish (**Figure 3A**). Among respondents who dropped out, the most frequently reported reason was lack of availability, accounting for 64.7% of responses. Additionally, 20.6% reported difficulty keeping pace with the course, and 8.8% perceived the course as too difficult. A smaller proportion of respondents (2.9%) were unable to complete the course due to hospitalization, while another 2.9% reported difficulties in submitting the required assignments (**Figure 3B**). Thus, time constraints rather than academic difficulty *per se* emerged as the primary barrier to completion, followed by challenges in maintaining course pacing.

**Figure 3:**
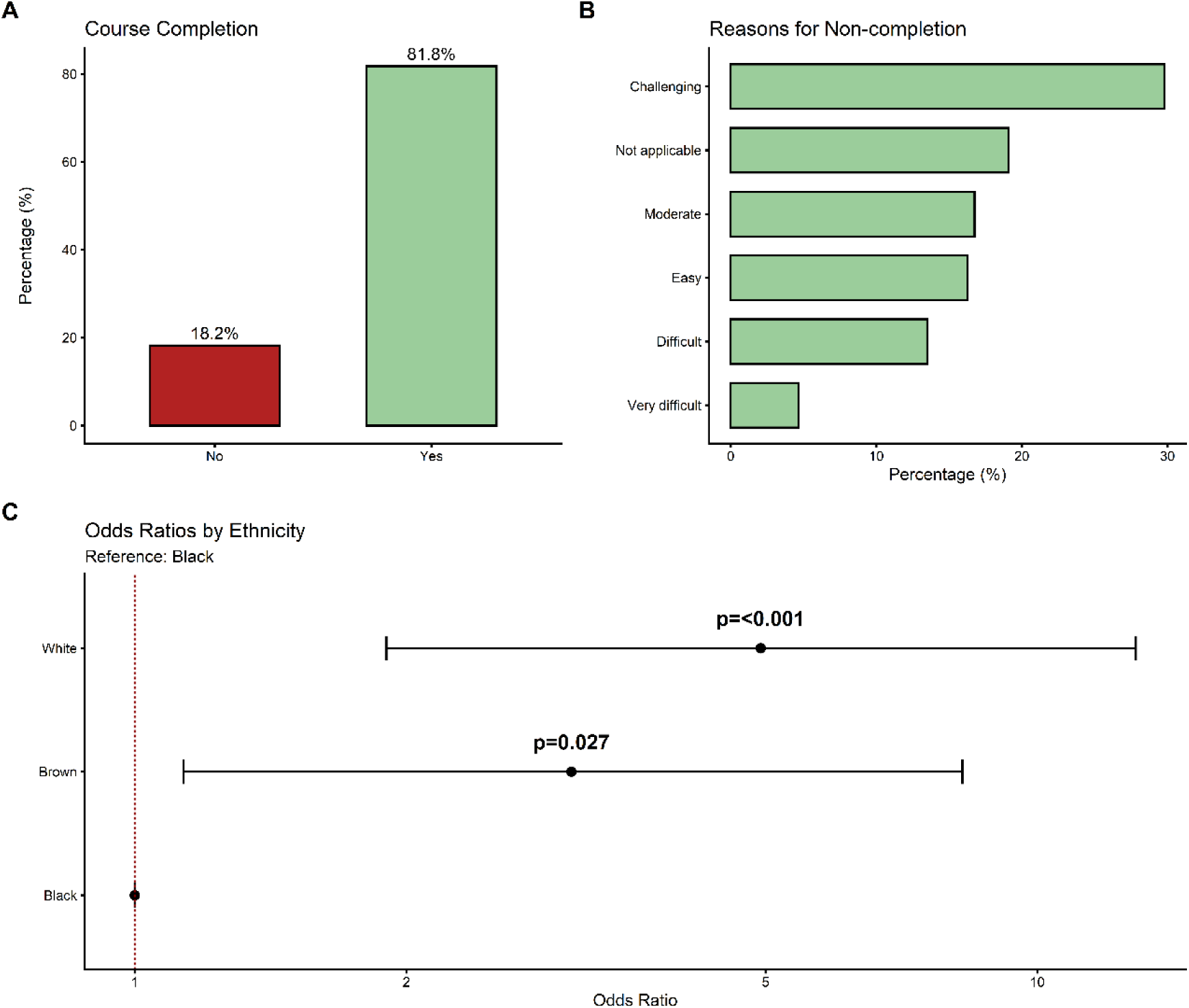
Analysis of course completion and reasons for non-completion by ethnicity. **(A)** Overall course completion rate among the 217 respondents. **(B)** Reasons for non-completion among those who did not finish the course. **(C)** Logistic regression results showing the likelihood of course completion by ethnicity, with the Black group as the reference (Logistic regression, *p*=0.0268 for Brown *vs* Black, *p*=0.00097 for White *vs* Black). Odds ratios: Brown OR 3.05, White OR 4.95.

To examine whether ethnicity predicted course completion, we performed a logistic regression analysis with the Black group as the reference category. Respondents of Brown ethnicity were (odds ratio [OR] 3.05) times more likely to complete the course than Black respondents (*p*=0.0268). White respondents showed an even larger advantage, being 4.95 times more likely to complete than Black respondents (OR 4.95; *p*=0.00097). These findings reveal a striking ethnic gradient in course success, favouring White and Brown individuals over Black individuals (**Figure 3C**).

Perceived difficulty also varied by ethnicity, though differences did not reach statistical significance (**Figure 4C**). The majority of respondents rated the course as “Challenging” (29.8%) or “Difficult” (19.1%), with only 4.7% selecting “Very Difficult” (**Figure 4A, 4B**). A violin plot examining the predicted probability of difficulty by ethnicity showed that White and “Other” ethnic groups had the highest predicted probabilities, whereas Black respondents had the lowest predicted probability of perceiving the course as difficult (*p*=0.0893, logistic regression; **Figure 4C**). Although this p-value exceeds the conventional threshold of 0.05, the trend suggests that Black respondents may have perceived the course as less difficult or were less likely to report difficulty a pattern that warrants further investigation in larger, more diverse samples.

**Figure 4:**
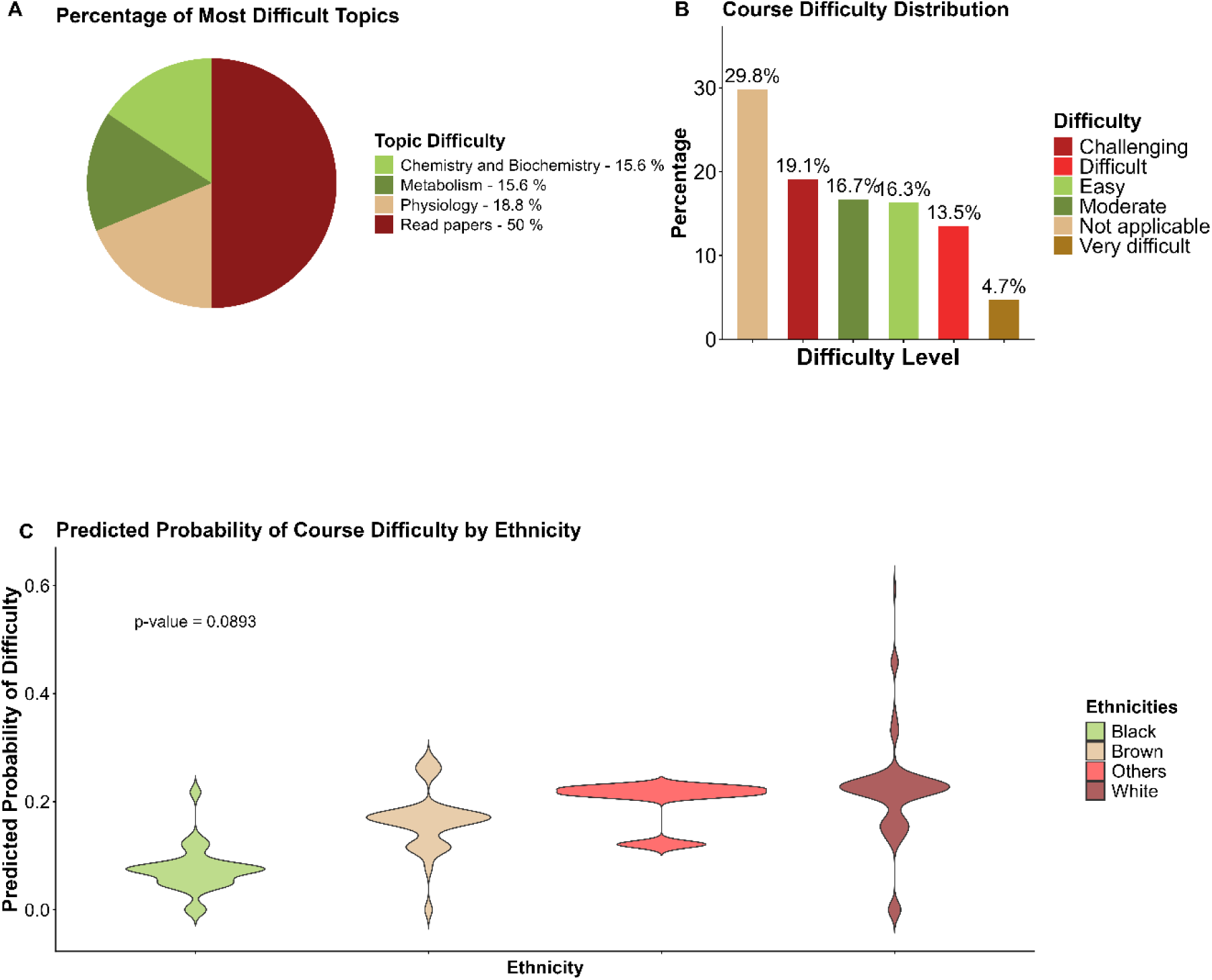
Distribution of course difficulty, most challenging topics, and predicted difficulty by ethnicity. **(A)** Proportional distribution of perceived course difficulty levels **(B)** Frequency of reported difficulty levels and most challenging topics **(C)** Predicted probability of course difficulty by ethnicity; logistic regression, *p*=0.0893. The violin plot shows predicted probabilities, with White and “Other” groups exhibiting higher values than the Black group.

Regarding specific challenging topics, Scientific Contents was the most frequently cited difficulty, accounting for 50% of all responses; this category specifically reflected challenges in reading and interpreting scientific texts and articles. In contrast, Chemistry and Biochemistry and Metabolism were each rated as difficult by 15.6% of respondents (**Figure 4B**).

### 3.3. From course completion to classroom practice: translating teacher learning into student health promotion

A central goal of the *Metabolism and Obesity* course was to equip teachers with practical, didactic-pedagogical activities that could be directly applied or adapted for use with their students. Throughout the course, at the end of each content block, a structured activity was suggested, allowing teachers either to implement it as designed or to formulate new activities based on the principles learned. These activities were proposed by the *Bio Cientista Mirim* program. To determine whether course completion translated into classroom practice, we examined the association between completion status and the self-reported application of these activities.

The chi-square test revealed a highly significant association between course completion and the application of learned activities in teaching (χ² test, *p*<2.2e-16; **Figure 5A**). Among respondents who completed the course, 57.1% reported having applied at least one activity in their classrooms, whereas 42.9% did not. By contrast, among those who did not complete the course, only 40% applied the activities, while 60% did not implement them. These findings indicate that course completion is associated with a substantially higher likelihood of translating educational content into teaching practice. Completion thus emerges not merely as a metric of individual persistence but as a critical enabler of knowledge dissemination to school-aged populations.

**Figure 5:**
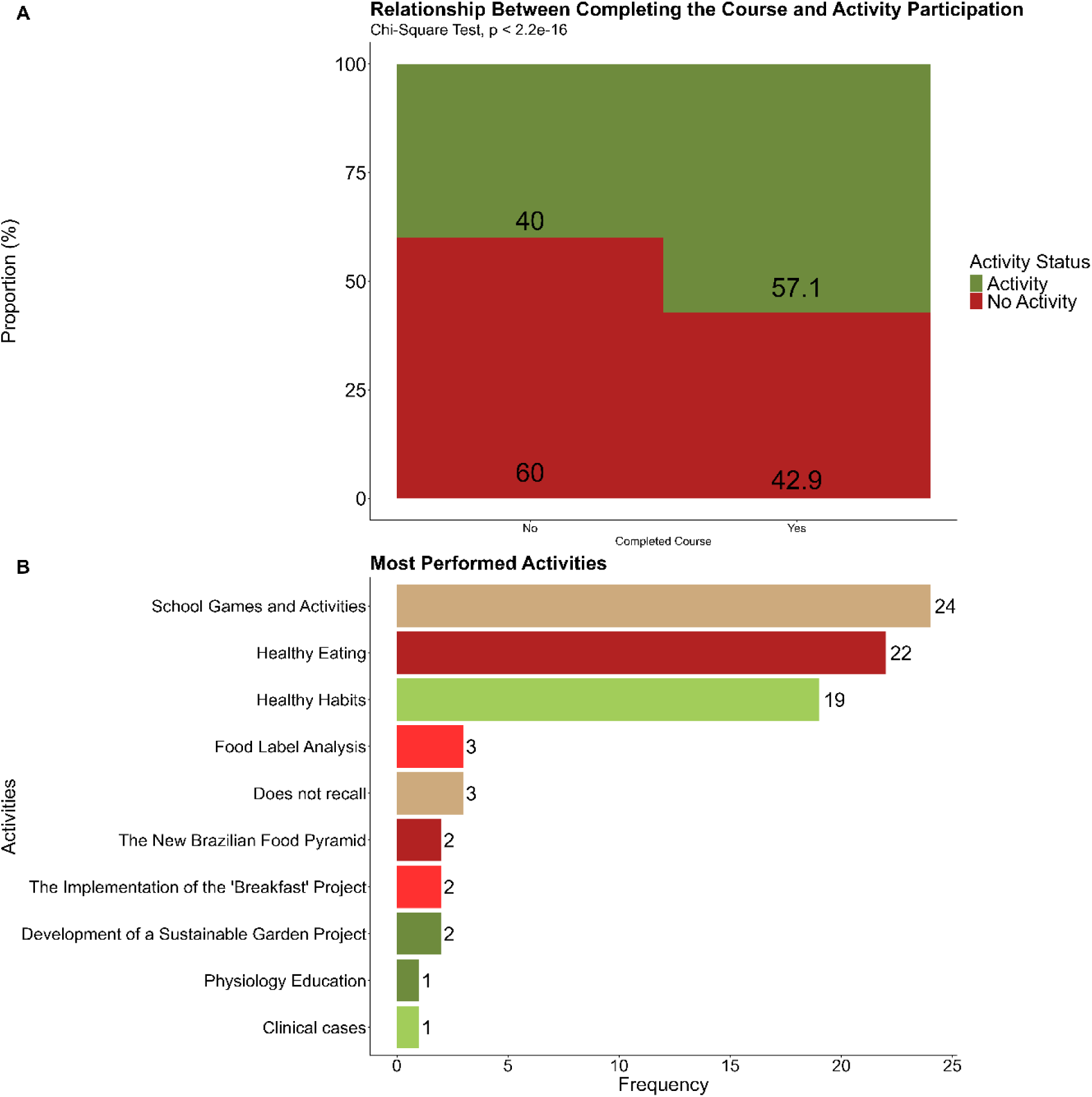
Relationship between course completion and participation in health promotion activities: frequency and types of activities performed. **(A)** Association between course completion and self-reported application of learned didactic-pedagogical activities in the classroom. Data are presented as percentages of respondents within each completion group. Statistical significance was assessed using the *chi-square* test (χ², *p*<2.2 x 10^-16^). **(B)** Frequency distribution of specific types of health promotion activities implemented by teachers in schools. Categories include Games and School Activities (n=24), Healthy Eating (n=22), Healthy Habits (diet plus physical exercise, n=19), Healthy Recipes (n=8), Dance and Physical Activity (n=6), and Others (n=5). Multiple responses were permitted.

We next characterised which types of health-promoting activities teachers most frequently implemented. As shown in **Figure 5B**, the most commonly reported activities were playful in nature: “School games and Activities” were cited 24 times, underscoring the popularity and perceived effectiveness of gamification in engaging students with health-related topics. “Healthy Eating” followed closely, mentioned 22 times, reflecting respondents’ strong emphasis on nutritional education and the importance of making informed food choices. “Healthy Habits” a category combining dietary practices with physical exercise, was recorded 19 times, indicating that teachers favoured a holistic approach to health that integrates behaviour change across multiple domains.

### 3.4. Course completion, lifestyle decisions and healthy habits: beyond the classroom

To assess the translational reach of the program beyond pedagogical application, we evaluated the intervention’s influence on the respondents’ reported changes in personal lifestyle choices and health-related habits. Our findings suggest that the course completation was associated with perceived behavioural chances, with robust statistical evidence supporting improvements in both lifestyle decisions and the adoption of health-promoting routines (**Figure 6**).

**Figure 6:**
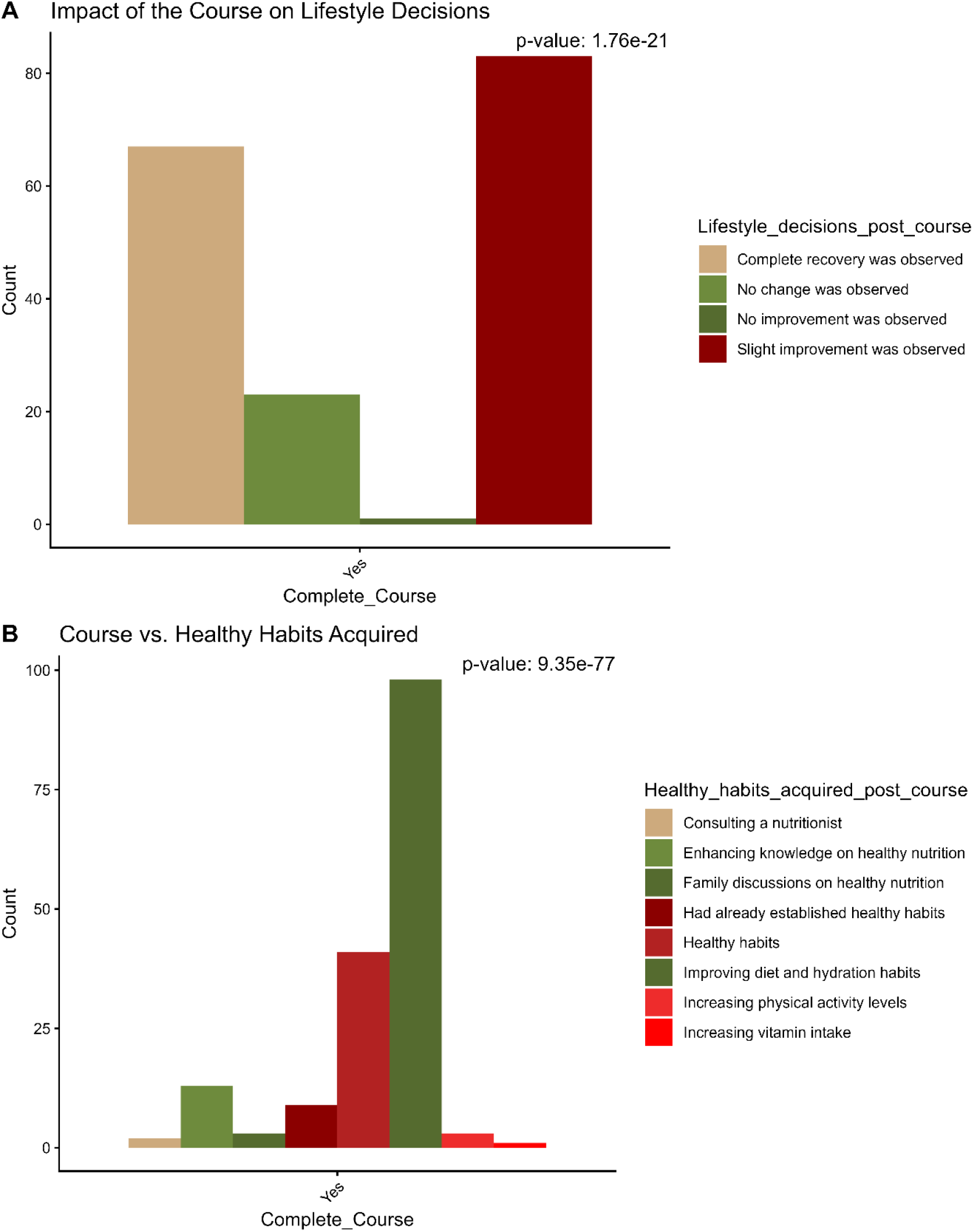
Impact of course completion on lifestyle decisions and the adoption of healthy habits. **(A)**. Association between course completion and self-reported lifestyle changes post-course (*chi-square* test, *p*=1.76 x 10^-21^). Response categories included “Complete recovery,” “Slight improvement,” “No improvement,” and “No change.” **(B)** Association between course completion and the adoption of specific healthy habits (*chi-square* test, *p*=9.35 x 10^-77^). Categories include “Enhancing knowledge on healthy nutrition,” “Consulting a nutritionist,” “Improving diet and hydration,” “Increasing physical activity levels,” and “Already had healthy habits prior to the course.” Multiple responses were permitted.

The vast majority of respondents who completed the curriculum/program reported substantial positive shifts in their lifestyle, ranging from “Slight improvement” to “Complete recovery” of healthy habits. Notably, cases of “No change” or “No improvement” were negligible among those who finalized the program. A chi-square analysis confirmed a highly significant association between course completion and improved lifestyle decisions (X*^2test^*, *p* = 1.76 x 10^-21^; **Figure 6A**), underscoring the program’s efficacy in fostering personal health agency.

Furthermore, completion of the intervention was strongly associated with the uptake of specific evidence-based healthy habits. Respondents reported significant increases in health literacy regarding nutrition, proactive dietary and hydration adjustments, and heightened physical activity levels, with some individuals seeking professional nutritional counselling following the course. The statistical significance of this association was exceptionally high; (X*^2test^*, *p* = 9.35 x 10^-77^; **Figure 6B**) confirming that the course completation not only reinforced pre-existing healthy behaviors but also served as a primary driver for the acquisition of new, health-protective habits. These results suggest that by educating the educator, the program generates a ripple effect of health promotion that transcends the classroom.

## 4 DISCUSSION

In this study, we found that a free, fully online course on metabolism and obesity reached and trained teachers across Brazil and was associated with positive reported outcomes, while also revealing persistent ethnic and regional disparities in access and completion. Our findings suggest that course completion is not merely an individual academic milestone but an important marker of knowledge translation into classroom practice and of positive lifestyle changes among educators themselves.

These findings align with the syndemic model of obesity proposed by Swinburn et al., (2019)^2^, in which schools represent strategic environments for intervention capable of simultaneously addressing obesity, undernutrition, and climate change through educational approaches^2^. The high engagement we observed, particularly the progressive increase in course participation beginning in 2021, reaching a peak growth rate of 27.6% in 2022, suggests that teachers recognize the relevance of metabolism and obesity content for their professional practice. This timing coincided with heightened health awareness following the COVID-19 pandemic, reinforcing that educational interventions can capitalize on windows of opportunity when health is at the forefront of public consciousness.

Extending this reasoning to classroom practice, our finding that 57.1% of course completers applied learned activities in their classrooms directly supports the assertion by Story et al., (1999) ^14^ that schools are strategic environments for health promotion, with teachers acting as mediators of knowledge and facilitators of behavioural change ^14^. The types of activities most frequently implemented, Games and School Activities (n=24), Healthy Eating (n=22), and Healthy Habits combining diet with physical exercise (n=19), reflect a holistic approach consistent with the syndemic perspective, which emphasises interconnected health behaviours rather than isolated risk factors ^2^.

Beyond these qualitative successes, the overall completion rate of 81.98% substantially exceeds the average completion rate reported for Massive Open Online Courses (MOOCs) globally, which typically ranges from 5% to 15% ^39,40^. This discrepancy is particularly notable when compared with Brazilian MOOC platforms such as Aprenda Mais, where completion rates average approximately 42.14% ^41^. The higher completion rate in our study probably reflects several factors: first, the course was offered specifically to educators with prior interest in metabolism and obesity, representing a motivated subset of the broader teacher population; second, the integration of structured didactic-pedagogical activities at the end of each content block enhanced engagement and provided immediate practical value; and third, the self-paced, asynchronous format accommodated teachers’ demanding schedules better than fixed-schedule courses. These findings align with those of Kizilcec et al., (2020)^32^, who demonstrated that self-regulated learning strategies and perceived relevance of course content are strong predictors of MOOC completion^32^.

However, high overall completion rates can mask important inequities. Our finding that White respondents were 4.95 times (*p*=0.00097) and Brown respondents 3.05 times (*p*=0.0268) more likely to complete the course than Black, reveals a striking ethnic gradient in online course success. These results are particularly concerning because Brazil’s population is majority non-White, approximately 56% Black or Brown according to the Brazilian Institute of Geography and Statistics ^42^, yet our sample was predominantly White (53.95%). This disparity persists even in an online environment often presumed to democratise access to education. Our findings align with recent evidence from Brazilian online education platforms, where demographic factors, notably race and colour, have been shown to significantly influence enrollment and completion rates ^43^. The reasons for these disparities are probably multifactorial. In this context, the importance of racial quotas, as established by the Brazilian Quota Law (Law 12.711/2012) in university settings, must be extended to other extension courses, promoting greater democratization and preventing free online courses from inadvertently reproducing the very inequalities they aim to reduce.

The predominance of respondents from São Paulo should be interpreted in light of the course dissemination strategy over time. Between 2017 and 2019, the course was disseminated through a formal collaboration with EFAP (Escola de Formação e Aperfeiçoamento dos Profissionais da Educação), which promoted participation among public school teachers in the state of São Paulo. In this context, course completion was recognised for career progression, providing a strong institutional incentive for enrolment. During the COVID-19 pandemic (2020–2021), the course became fully accessible nationwide, facilitating participation from teachers across Brazil and increasing geographical diversity among respondents. Nevertheless, São Paulo remained the predominant state throughout the study period, suggesting that institutional dissemination channels and professional incentives may have contributed to the concentration of participants in this region. Therefore, the observed geographical imbalance should be interpreted in the context of the programme’s dissemination strategy. A limitation of this study is the overrepresentation of participants from São Paulo, which may have influenced participant distribution and limits the generalisability of findings to the broader Brazilian population.

Kucirkova (2023)^44^ have documented that digital inequality extends beyond mere access to devices and connectivity, encompassing differences in digital literacy, social support networks, and culturally responsive content design^44^. Black and Brown teachers in Brazil may face additional structural barriers, including higher teaching loads, fewer planning hours, and greater exposure to under-resourced schools, all of which can limit the time and cognitive energy available for professional development ^45–47^. Moreover, research shows that Black students often work harder to prove themselves due to racist stereotypes, yet this high effort carries emotional costs when institutions do not address underlying racism ^48,49^. Our finding that Black respondents who persisted rated the course as less difficult may reflect this phenomenon.

Adding nuance to this disparity, the trend we observed for perceived difficulty (*p*=0.0893) suggests that Black respondents had the lowest predicted probability of rating the course as difficult. Although this finding did not reach statistical significance, it raises the possibility that Black teachers who persisted in the course may have been a highly selected group, those with the strongest prior knowledge or motivation, or alternatively, that measurement of perceived difficulty may have been influenced by cultural differences in response styles. Future qualitative research is urgently needed to explore the barriers and facilitators experienced by Black and Brown teachers in online professional development.

Turning from disparities to outcomes of success, our finding that course completion was strongly associated with application of learned activities (χ² test, p<2.2 x 10^-16^) adds to the growing evidence base supporting school-based health education interventions. A systematic review and meta-analysis by Jacob et al., (2021) ^19^ concluded that multi-component school-based interventions delivered by trained teachers can significantly reduce BMI z-score, albeit by a modest margin (−0.06 [95% CI −0.10, −0.03])^19^. In parallel, a growing body of research indicates that teacher-delivered, multi-component approaches, which integrate educational content with environmental modifications and, in many cases, digital tools or parental engagement, yield small yet consistent benefits across reviews and clinical trials ^50^. Importantly, these reviews identified teacher training as a key feature of effective interventions, along with parental involvement and digital components. Our study extends these findings by demonstrating that online-only teacher training, without in-person component, can still produce meaningful changes in teacher behavior, suggesting that scalable digital interventions may offer a cost-effective complement to face-to-face professional development.

The types of activities teachers chose to implement further reinforce this point. The most frequently implemented activities in our study, Games and School Activities, Healthy Eating, and Healthy Habits are consistent with evidence-based strategies for engaging students. A systematic review of school-based nutrition interventions found that interactive, participatory methods (including games, cooking demonstrations, and hands-on activities) produce larger improvements in dietary knowledge and behaviours than do didactic approaches alone ^16^. Murimi et al., (2018)^51^ found that nutrition programmes are more likely to succeed when they include experiential elements like gardening, gaming, and cooking. Similarly, Charlton et al., (2020)^52^ reported that school gardens, cooking classes, taste testing, and multi-component programmes help improve children’s willingness to try new foods, their cooking skills, and their nutritional knowledge. Across age groups, hands-on, play-based interventions consistently outperform passive, instruction-only lessons, which tend to boost knowledge but rarely change eating habits^16^. Taken together, these findings suggest that teachers intuitively recognise what works, favouring playful, practical activities over more conventional teaching methods.

Perhaps most notably, the benefits of course completion extended beyond the classroom into teachers’ personal lives. The finding that course completion was associated with significant improvements in teachers’ own lifestyle decisions (*p*=1.76 x 10^-21^) and adoption of healthy habits (*p*=9.35 x 10^-77^) is noteworthy for two reasons. First, it addresses a gap in the literature identified by Hill et al., (2022)^26^, who noted that teacher well-being is often overlooked in school-based health interventions, despite evidence that teachers’ personal health behaviours influence their teaching practices and their ability to act as role models^26^. Our results suggest that structured online courses that integrate practical, applicable content can serve as catalysts for personal behaviour change, probably because teachers who internalise health messages for themselves are better equipped to model and teach them to students. Second, this finding aligns with the teacher-as-role-model effect described in school-based health interventions^53^. When teachers adopt healthier behaviours improving diet, increasing physical activity, or consulting a nutritionist, they not only experience direct health benefits but also model these behaviours for students, potentially amplifying the intervention’s impact beyond the classroom. This win-win outcome is particularly valuable in resource-limited settings, where every investment in professional development should ideally yield multiple benefits.

Nevertheless, not all content areas were equally accessible. The most challenging topics reported by respondents, Scientific Content/read papers (50% of responses), followed by Chemistry and Biochemistry, and Metabolism (15.6% each), point to specific areas where course design could be improved. These findings are consistent with the broader literature on teacher professional development in health education, which has shown that teachers often struggle to translate complex biomedical concepts into age-appropriate, curriculum-aligned lessons ^54,55^. This difficulty likely reflects a broader gap in teachers’ undergraduate preparation, as many Brazilian educators have limited exposure to advanced concepts in metabolism, biochemistry, and physiology during their initial training ^56,57^. As one potential way forward, we are developing computer games designed to help teachers overcome this difficulty. A natural next step will be to evaluate the effectiveness of these games, both with teachers themselves and within real classroom environments. Future iterations of the course could also benefit from additional scaffolding, such as glossaries, visual aids, ready-to-use classroom materials, and optional deep-dive modules for those seeking deeper understanding without overwhelming the core curriculum.

Compounding these content-specific challenges are broader structural barriers to access. The geographical concentration of respondents in São Paulo (77%) and the complete absence of respondents from several northern states (Acre, Roraima, and Amapá) highlight persistent digital divides in Brazil. These findings are consistent with national data on internet access, which show that northern and northeastern regions have lower rates of broadband connectivity and lower digital literacy than the southeast ^37,58^. Importantly, the course was offered entirely online, with no offline or hybrid alternatives. Although web-based education enables flexible, anywhere, anytime access^28^, this flexibility is only meaningful when reliable internet infrastructure exists. Teachers in remote or resource-poor regions may face connectivity issues, lack of access to appropriate devices, or electricity instability that precludes consistent participation ^37^. Future scaling strategies should consider offering hybrid options, including downloadable content that can be accessed offline, mobile-optimised versions for low-bandwidth settings, and community-based study groups with shared internet access.

A key implication of this study is the warning that ‘scalability without equity’ is a pitfall for public health policies. While the program successfully translated complex scientific concepts for teachers, the ethnic and regional completion gaps indicate that future digital health models must incorporate specific strategies to support vulnerable groups. True ‘Translational Pedagogy’ must be inclusive, ensuring that technological advancement does not exacerbate social injustices. Several limitations warrant consideration: the cross-section post-course design precludes causal inference; outcomes were self-reported, raising social desirability bias; the small number of Black respondents (n=27) and geographical imbalances (77% from São Paulo) limit generalisability; and we did not measure student-level outcomes or confounders such as socioeconomic status and technology access. Despite these limitations, this study makes important contributions as the first systematic evaluation of a web-based Metabolism and Obesity course for Brazilian teachers, suggesting that online-only training can produce meaningful changes in both classroom practice and teachers’ personal lifestyles.

## 5 CONCLUSIONS

This study suggests that completion of an online course on metabolism and obesity was associated with reported improvements in Brazilian teachers’ lifestyle behaviours, including diet, physical activity, and nutrition knowledge, as well as with classroom health promotion activities. However, marked ethnic and regional disparities in completion, with White teachers nearly five times more likely to complete the course than Black teachers and several northern states unrepresented, reveal critical equity gaps. Free and online access alone does not democratise education; digital inequality extends beyond connectivity.

Three priorities emerge: randomised controlled trials to establish causality with student-level outcomes, qualitative studies of barriers faced by Black and Brown teachers, and implementation science to identify supports that maximise completion and knowledge translation in underserved regions.

Online professional development is scalable but not inherently equitable. Targeted strategies, inclusive design, recruitment of underrepresented groups, and infrastructure investment, are essential. The challenge now is to ensure that scalable digital interventions reduce, rather than reproduce, health and educational inequities across Brazil’s diverse teaching workforce.

## Credit authorship contribution statement

**P.H.E-S**: Conceptualization, Methodology, Investigation, Visualization, Collected and analyzed data, Formal analysis, Writing – original draft, Writing – review & editing. **C.P.C**: Formal analysis, Methodology, Investigation, Visualization, Collected and analyzed data, Writing – original draft, Writing – review & editing. **C.C.M.C**: Formal analysis, Methodology, Investigation, Visualization, Collected and analyzed data, Review & editing. **T.M.S**: Investigation and Methodology, and Writing – review & editing. **A.F.F.F**: Investigation, and Methodology, and Writing – review & editing **A.C.M.K**: Investigation and Methodology. **J.K.N.R**: Investigation. **L.A**: Investigation **R.L.A.F**: Conceptualization, Investigation, and Methodology **C.R.O.C**: Conceptualization, Formal analysis, Writing – original draft, Writing – review & editing. All authors approved the final version of the manuscript and agree to be accountable for all aspects of the work.

## Declaration of Generative AI and AI-assisted technologies

The authors used DeepSeek solely for Portuguese-to-English translation and language polishing during manuscript preparation. No AI was used for data analysis, interpretation, conclusion drawing, or generation of scientific content. The authors reviewed all changes and assume full responsibility for the final work. This statement complies with The Lancet policies on generative AI.

## Data Availability

All data produced in the present study are available upon reasonable request to the authors

## Acknowledgments

The authors thank all participants from all editions of the Metabolism and Obesity course (2017–2022) who generously volunteered their time to complete the evaluation questionnaire and share their experiences. The authors also thank the professors and graduate students from Department of Physiology and Biophysics from ICB-USP, who delivered lectures and facilitated the course modules over the years, whose dedication and expertise were fundamental to the implementation of this educational intervention.

## Inclusion and diversity

One or more of the authors of this paper self-identifies as an underrepresented ethnic minority in their field of research or within their geographical location. One or more of the authors of this paper self-identifies as a member of the LGBTQIA+ community.

