## Supplementary material for "Web-based education on Metabolism and Obesity is associated with improved lifestyle and health behaviours among Brazilian school teachers": Suplementary material 1

**[#]Contributed equally**

### Supplementary material 1

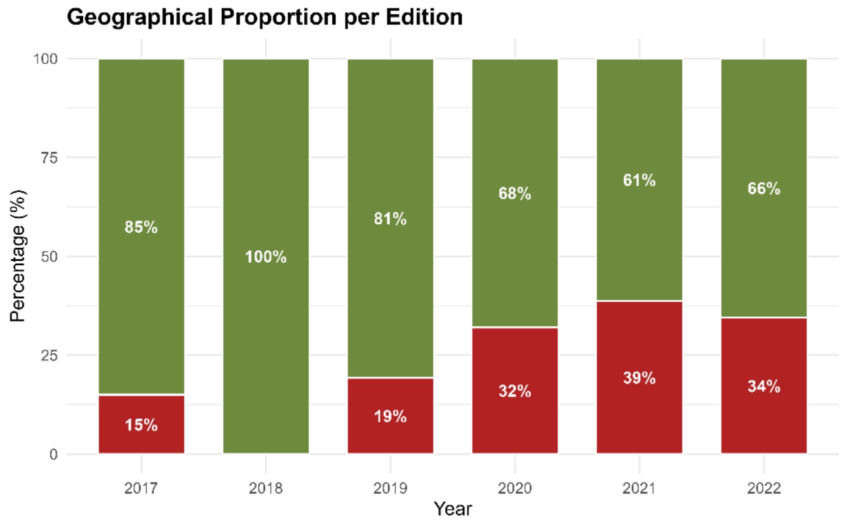


**Supplementary Figure S1 - Geographic distribution and expansion of responders across editions in Brazil. (A)** Geographic proportion of participants per edition. Each bar represents the relative distribution of participants among Brazilian regions or states across different editions. São Paulo consistently accounts for the largest share, although its relative proportion varies over time. Categories with negligible proportions are not labeled.
